# WHAT ABOUT USING SNIFFIN’ STICKS 12 SCREENING TEST TO IDENTIFY POST-COVID-19 OLFACTORY DISORDERS?

**DOI:** 10.1101/2021.06.06.21258430

**Authors:** Clair Vandersteen, Magali Payne, Louise-Émilie Dumas, Alexandra Plonka, Grégoire D’Andrea, David Chirio, Élisa Demonchy, Karine Risso, Florence Askenazy-Gittard, Nicolas Guevara, Laurent Castillo, Valeria Manera, Auriane Gros

## Abstract

**Background:** Olfactory impairment is a major sequela of SARS-CoV-2 infection and has a negative impact on daily life quality. Olfactory loss can be assessed in many ways but seems to be little realized in a daily clinical practice. The sniffin Sticks test – 12 items (SST-12) can be used in quick olfactory disorders screening. Its use in a post-covid19 situation was the main objective of this work.

**Methodology:** Consecutive patients consulting to the ENT department with post-Covid-19 olfactory loss were included. The clinical examination included an analog scale for the self-assessment of olfactory recovery (VAS), self-reported salt and sugar intake, a nasofibroscopy, the complete Sniffin’ Stick Test (SST) and the SST-12.

**Results:** Among the 54 patients included, based on the SST-12, 14,8% (n=8) of the patients could be classified as normosmic (SST-12≥11), 48,1% (n=26) as hyposmic (6< SST-12<10) and 37% (n=20) as functional anosmic (SST-12≤6). We report excellent and significant correlations between SST-12 and SST or VAS assessments. Salt and Sugar increased intake seems significantly related to SST-12 results.

**Conclusions:** SST-12 is a reliable way to screen post-COVID-19 olfactory disorders could be used in a daily clinical practice and might be used to prevent bad diet habits and so cardiovascular risk.

## INTRODUCTION

The onset of a sudden partial (hyposmia) or total (anosmia) loss of smell is now recognized as highly predictive of SARS-COV-2 infection^(1)^. Anosmia can be the only COVID-19 symptom in 11 to 26% of cases^(2–4)^. The long-term anosmia can cause an alteration in the quality of life^(5)^ and psychiatric disorders such as depression^(6,7)^, anxiety, anorexia^(8)^ and its nutritional consequences^(9)^, social interaction disorders^(10,11)^ and cognitive impairment^(10,12,13)^. So, the diagnosis of olfactory disorders and their management is essential especially as 6^(14)^ and 12^(16)^ months after the COVID-19 infection, respectively 60% and 30% of patients retain an olfactory complaint and require attention. Moreover, even if sweet, salty, sour and bitter tastes improved from 60% (acute COVID-19) to 97.2% (6 months) of patients^(14)^, salt and sugar intake increase concerned near 30% of post-COVID-19 patients^(15)^, especially young women. These findings should require patient’s education to prevent cardiovascular risk.

Although there are different ways to assess a patient’s ortho and retro-olfaction ^(17,18)^, only 50% of ENTs assess the olfactory disorders on an anamnesis, and 10% assess through psychophysical tests ^(19)^. Olfaction is most often evaluated by subjective self / hetero questionnaires with a significant variability of the results and a probable underestimation,^(19,20)^ given the poorer olfactory perception before 20 years and after 50 ^(21,22)^. Complete psychophysical olfactory tests, with assessment of odor threshold, odor discrimination and odor identification, are the gold standard ^(19)^ and allow to specify the olfactory disorder ^(23)^. The most used in Europe is the Sniffin ‘stick test® (SST)^(19,21,24–26)^ that include an odor Threshold detection (T), an odor Discrimination (D) and an odor Identification (I) tests. However, these psychophysical tests are expensive and take a long time (between 30 and 60 minutes))^(17,18)^, thus making their daily clinical use difficult. It therefore seems important to look for other olfactory tests that are faster (≤5 minutes) and accessible to specialists, but also to general practitioners.

The Sniffin ‘Sticks Test - 12 items (SST-12) is an olfactory screening test in the form of a 4-minute identification test allowing, according to its authors, to detect anosmia and hyposmia with comparable measurement reliability other similar scent screening tests ^(27,28)^. It can also be used laterally (one nostril tested independently of the other).

Seeing that it has been demonstrated that identification disorders are predominant in post COVIDs ^(5)^, the objective of this study was to assess the value of SST-12 in the detection and characterization of a persistent post-COVID-19 olfactory disorder.

## MATERIAL AND METHODS

### Population

The study was approved by the institutional review board of the Nice University Hospital (CNIL number: 412). This study is part of a large work registered under a ClinicalTrials.gov number (ID: NCT04799977). Since March 2020, we retroprospectively recruited at ENT department of Nice University Hospital all patients infected by COVID-19 with persistent olfactory disorders from two to nine months. Patients where self-referred or referred by colleagues, general practitioners or advised by the infectiology department that managed all COVID-19 declared patients (city guidelines). Patients had either an olfactory complaint for over 6 weeks and a molecular-proven SARS-CoV-2 diagnosis or a CT-proven SARS-CoV-2 diagnosis secondarily confirmed by serology. We retrospectively extracted patients’ demographic data, and clinical characteristics including nasofibroscopy, visual analogue scale (VAS) for the subjective assessment of olfactory recovery (ranging from 0% to 100%), subjective taste impairment, over intake of salt and sugar, and SST^(26,29,30)^ total and subdomains results which were systematically assessed. SST-12 results were extrapolated from SST results.

### Sniffin’ sticks test 12 items

Olfaction diseases SST-12 test has been validated in 2001 by Hummel et al.^(31)^. This 4 min screening psychophysical test is an odor identification test based on 12 from the 16 odors being sniffed during the identification subdomain part of the original SST.

The original SST identification odors set include peppermint, orange, fish, leather, rose, cloves, coffee, pineapple, licorice, anise, lemon, banana, cinnamon, apple, turpentine and garlic. During the identification SST test, subjects were blindfolded. Sixteen odorant sticks were presented once, separated by an interval of at least 20 seconds to prevent olfactory desensitization. Each stick presentation was accompanied by a written list containing the correct odorant and 3 semantic distractors. Retrospectively, results from all odors set but apple, turpentine, garlic and anise were summed up to the SST-12 global score, as previously described^(31)^. We defined a normosmia (SST-12≥11), an hyposmia (10>SST-12>6) or an anosmia (SST-12≤6) based on normative values assessed from more than 1200 patients assessed with SST and olfactive evoked potential for anosmic and hyposmic ones^(31)^. Apple, turpentine and garlic have been removed from the SST-12 because identified by less than 55% of its normosmic validation cohort^(31)^. Anise was removed too because of being too similar to liquorice. With a reproducibility kappa coefficient of 0,77, the diagnosis agreement can be considered as “good” (Altman, 1991). Although olfactory abilities decreased at extreme ages, SST-12 can be used before the age of 10 and after the age of 80.

### Statistical Analysis

Data are presented as mean (SD) for quantitative variables and as frequency and percentage for qualitative variables. The degree of accordance between the SST and the SST-12 in patients’ categorization was calculated using Cohen’s Kappa coefficient. Sensitivity and specificity of the SST-12 compared to the SST in classifying patients as anosmic, was also reported. To verify whether patients that increased their consumption of salt and sugar had lower SST and SST-12 scores compared to those who did not, we employed Mann-Whitney U tests. Chi2 tests were employed to explore links between self-reported taste disorders and the presence of an increased salt and sugar consumption. To investigate correlations between subjective reports (VAS), and odor identification disorders (based on the SST and SST-12) we performed bivariate correlation analyses. As data were not normally distributed (as suggested by Kolmogorov-Smirnov test), non-parametric Spearman’s correlations were employed.

## RESULTS

### Demographic and clinical features

Fifty-four patients consulting the ENT department of Nice University Hospitals (CHU) for olfactory complaints after a COVID-19 infection were included in the study. The demographic and clinical features are reported in Table 1. 57% of patients were female (n=31), with a mean age of 39.9±13,9 years. They were seen after 5.4±3,1 months after the COVID-19 infection. 17 patients (31.5%) received a COVID-19 related treatment.

**Table 1.** Demographic and clinical characteristics. SD=standard deviation; CT=computerized tomography; PCR=polymerase chain reaction

|  | n | % |
| --- | --- | --- |
| **Total** | 54 | 100 |
| **COVID<sub>19</sub> testing** |  |  |
| Molecular PCR test | 46 | 85.2 |
| Chest CT | 11 | 20.4 |
| Serology (antibody test) | 16 | 29.6 |
| **COVID-19 dedicated treatment** |  |  |
| Oral corticosteroids | 6 | 11.1 |
| Nasal corticosteroids | 4 | 7.4 |
| Inhaled corticosteroids | 2 | 3.7 |
| Azithromycin alone | 7 | 13.0 |
| Hydroxychloroquine alone | 1 | 1.9 |
| Azithromycin + Hydroxychloroquine | 3 | 5.5 |
| Amoxicillin alone | 1 | 1.9 |
| Amoxicillin + Azithromycin | 2 | 3.7 |
| Others (vitamins, zinc) | 5 | 9.3 |
|  | mean | SD |
| **VAS (subjective % of olfactory recovery)** | 33.9 | 25.6 |
| **Sniffin' Sticks test – scores** |  |  |
| Threshold detection | 4.7 | 4.0 |
| Discrimination | 10.3 | 3.1 |
| Identification | 9.4 | 3.9 |
| **Taste disorders** | 49 | 90.7 |
| Retro-olfaction alone | 35 | 64.8 |
| Retro-olfaction + taste | 12 | 22.2 |
| Taste alone | 1 | 1.8 |

### Retrospective olfactory and taste complains screening results

Descriptive analyses for the loss of smell and taste are reported in Table 2. The day of consultation, patients reported to have recovered only 33.9±25.6% of their olfaction (ranging from 0% to 90%). 90.7% of the patients (n=49) reported taste disorders, including retro-olfaction (food flavors) alone (64.8%, n=35), retro-olfaction associated to taste (22.2%, n=12; 16.7% concerning sweet and salty, 11.1% concerning sour and bitter), or taste alone (1.8%, n=1 concerning sweet and salty). 45.5% of patients (20 out of the 44 who responded to the question) reported that they increased their consummation of salt, and 20.5% (9 out of 44) that they increased their consummation of sugar.

**Table 2.** Categorization of subjects based on the Sniffin’ Sticks Test scores (SST), Sniffin’ Sticks Test 12 items scores (SST12) and inter-test reliability (Kappa)

| **N=54** | **Normosmic** | **Hyposmic** | **Anosmic** |
| --- | --- | --- | --- |
|  | **N(%)** | **N(%)** | **N(%)** |
| **SST** | 13(24,1) | 29(53,7) | 12(22,2) |
| **SST-12** | 8(14,8) | 26(48,1) | 20(37,0) |
| *Correct Categorization* | 4(7,4) | 17(31,5) | 12(22,2) |
| *False positive* | 0(0) | 9(16,7) | 8(14,8) |
| *False negative* | 4(7,4) | 0(0) | 0(0) |
| **Cohen’s Kappa** | 0,242 | 0,224 | 0,654 |

Categorization of patients based on the results of the olfactory tests is presented in Table 2. The global results (TDI) of the Sniffin’ Sticks Test (SST) suggested that 24,1% (n=13) of the patients could be classified as normosmic (TDI≥30.75), 53,7% (n=29) as hyposmic (16.25≤TDI≤30.5) and 22,2% (n=12) as functional anosmic (TDI≤16). Based on the SST-12, 14,8% (n=8) of the patients could be classified as normosmic (SST-12≥11), 48,1% (n=26) as hyposmic (6< SST-12<10) and 37% (n=20) as functional anosmic (SST-12≤6). Interestingly, patients that increased their consummation of salt showed lower SST (U=112.5, p=0.003) and SST-12 (U=121, p=0.005) scored compared to the patients that did not increase salt usage (20.0±8.8 vs. 27.9±7.7, and 5.7±3.5 vs. 8.5±2.2, respectively). The same result was found for patients who increased their consummation of sugar, that showed lower SST (U=73.5, p=0.014) and SST-12 (U=62, p=0.005) scored compared to the other patients (26.2±8.0 vs. 17.1±8.4, and 4.6±2.9 vs. 6.9±2.9, respectively). The self-reported presence of taste disorders did not show any significant link with the presence of an increased consummation of salt (Chi^2^=0.74, p=389) or sugar (Chi^2^=1.13, p=287).

Taking SST as the *gold standard*, on 54 patients, 61% (n=33) were classified in the same category by the SST-12 patients. SST-12 misdiagnosed 4 patients as normosmic (7.4%), 8 as anosmic (14.8%), and 9 as hyposmic (16.7%). Importantly, all the patients that were diagnosed as anosmic by the SST were also detected by the SST-12. Accordingly, Cohen’s Kappa coefficient revealed a week agreement between the two tests in classifying patients as normosmic (Kappa= 0.24) and hyposmic (Kappa=0.22), but a strong agreement in classifying patients as anosmic (Kappa=0.65). The sensitivity and specificity of the SST-12, compared to the SST score, is reported in Table 3 and suggests that a score of 6 is the cut-off that maximize the combination between specificity (100%) and sensitivity (81%) in detecting anosmic patients. The presence of taste disorders did not affect the type of errors of the SST-12 compared to the SST.

**Table 3.**
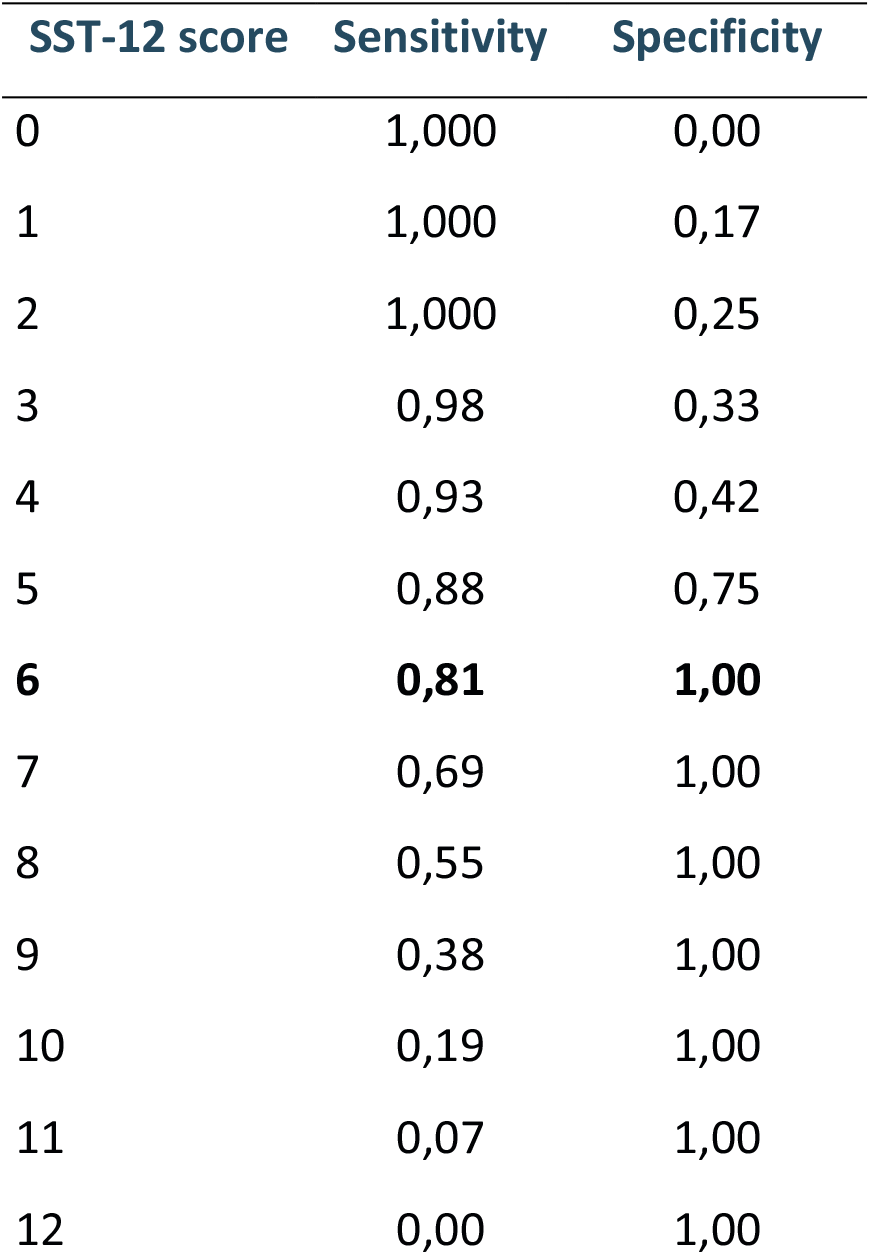
Sensitivity and specificity of the different cut-off scores of the SST-12 compared to the SST in classifying patients as anosmic

| SST-12 score | Sensitivity | Specificity |
| --- | --- | --- |
| 0 | 1,000 | 0,00 |
| 1 | 1,000 | 0,17 |
| 2 | 1,000 | 0,25 |
| 3 | 0,98 | 0,33 |
| 4 | 0,93 | 0,42 |
| 5 | 0,88 | 0,75 |
| **6** | **0,81** | **1,00** |
| 7 | 0,69 | 1,00 |
| 8 | 0,55 | 1,00 |
| 9 | 0,38 | 1,00 |
| 10 | 0,19 | 1,00 |
| 11 | 0,07 | 1,00 |
| 12 | 0,00 | 1,00 |

### Correlations between self-reported olfactory recovery, SST and SST-12 score

VAS scores were 45±24% (range 5%-70%), 38±25% (range 1%-90%), and 13±16% (range 0%-50%) for respectively normosmic, hyposmic and anosmic patients, based on the SST. Based on the SST-12, VAS scores were 52±26%, 42±22%, and 16±19% for respectively normosmic, hyposmic and anosmic patients. An almost perfect correlation between scores at the SST and SST-12 was found (rho_(52)_=0.98, p<0.001), confirming that the SST-12 can assess odor identification as well as the SST. Correlations between subjective reports (VAS) and the SST and SST-12 scores suggested a significant, positive correlation between percentage of subjective olfactory recovery (VAS) and the identifications scores for both the SST (rho_(52)_=0.47, p< 0.001) and the SST-12 scores (rho_(52)_=0.49, p< 0.001), testifying that the two scales were equally correlated to self-reported disorders. These results are reported in Figure 1.

**Figure 1.**
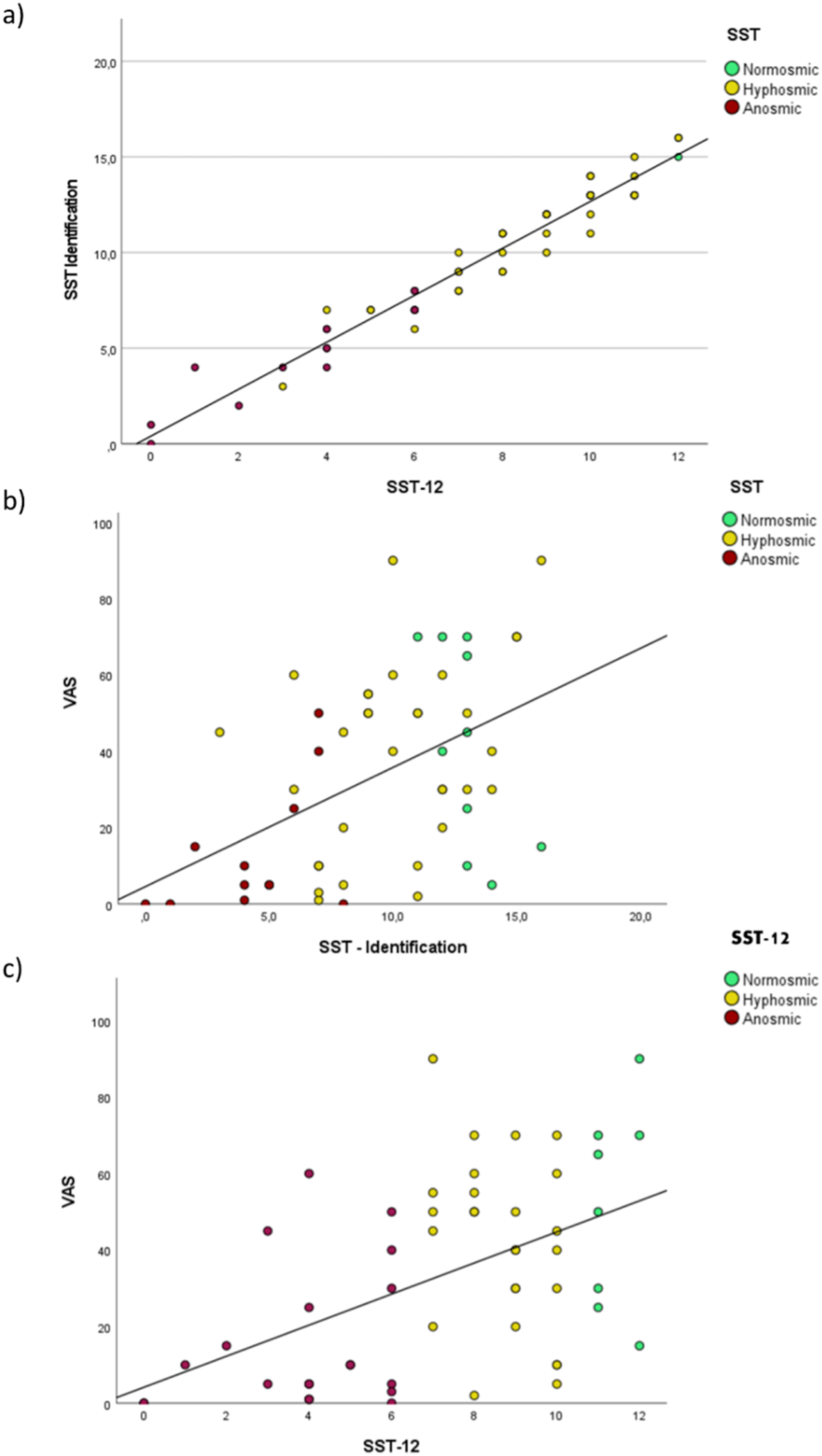
Correlations between a) SST-12 and SST identification score, b) percentage of subjective olfactory recovery (VAS) and SST, and c) percentage of subjective olfactory recovery (VAS) and SST-12

## DISCUSSION

This is the first study that evaluates quantitatively the efficiency of the SST-12 to screen for post-COVID-19 olfactory disorders, and specially to identify post COVID-19 anosmics patients.

Screening for olfactory disorders is important because, in addition to allowing to set up appropriate care for patients, it helps prevent the occurrence of consequences of long-term anosmia like an alteration in the quality of life^(5)^, psychiatric disorders such as depression^(6,7)^, anxiety, anorexia^(8)^ and its nutritional consequences^(9)^, social interaction disorders^(10,11)^ or cognitive impairment^(10,12,13)^. Although a subjective olfactory complaint (80% anosmia, 20% hyposmia) is now a very frequent symptom of a COVID-19 infection ^(2)^ affecting 70 to 85% of patients ^(32,33)^, only 21% of clinicians use psychophysical olfactory tests to characterize this olfactory complaint ^(20)^. Odor disorder is also the only symptom in 16-20% of cases (3,34).

Post-COVID-19 olfactory disorders show unique psychophysical characteristics in the long term. In a population of 34 patients deprived of their olfaction for about 6 months after COVID-19 and presenting a persistent odor complaint (VAS), we previously highlighted a significantly predominant impairment of odor identification^(5)^, characteristic of central olfactory impairment ^(23)^. This impairment worsened with the duration of olfactory deprivation and significantly impacted the quality of life.

The present study shows the reliability of SST-12 in screening for post-COVID-19 olfactory disorders, and in particular anosmia. Among SST-12 diagnostic errors, only 7% (n = 4) of hyposmic patients would have been considered normosmic by the gold standard (SST). The other differences in scores between the SST-12 and the SST do not modify the purpose of the screening, which is to perform or have performed complete olfactory tests in the event of an abnormality detected. In this way, all but 4 patients (92%) would have been correctly screened using the SST-12. All the anosmics patients at the SST were correctly screened by the SST-12 as evidenced by the “good” correlation coefficient (0,61≤Kappa≤0,80 - Altman 1991). Other screening test do exist. The Q-stick test^(35,36)^ is another olfactory screening test validated on 196 people from the SST. It allows, after the age of 12, to assess the identification of three smells (coffee, cloves and rose) contained again in olfactory markers. On the other hand, its reliability coefficient is less good than the SST-12 (Kappa between 0.2 and 0.33 at most)^(36)^. Also, it does not detect 8% of anosmics according to its authors^(36)^. Others are more used in Asia or the American continent (Cross Cultural Smell Identification Test (CC-SIT)^(27)^, Q-SIT^(28)^) but they are based on single-use “scratch and sniff”. Others are less used^(17)^ (Kremer olfactory Test, Le Nez du Vin, Smell Diskettes). They require additional specific equipment (Jet Stream Olfactometer) or have been validated outside of Europe (different scents) or on small cohorts, and never in the context of COVID-19.

The total SST-12, as SST, was significantly correlated (p <0.001) with odor complaint (VAS – figure 1) which reflects persistent post-COVID-19 olfactory impairment, i.e. identification disorder ^(5)^. Unlike the SST, the SST-12 only assesses the identification of odors and thus seems more suited to detect an identification disorder than the SST, which adds to the identification score, a score for discrimination and perception of the odor threshold. As we have shown in previous studies ^(5)^, the SST interpretation can conclude to a global normosmia when one of its subdomains is altered. The SST should therefore not be used as a gold standard in the post-COVID-19 odor evaluation given that some patients, early^(37)^ and at a distance^(5)^ from SARS-COV-2 infection, may be incorrectly classified as normosmic on SST despite odor loss^(5)^. In this study we found that olfactory complain (VAS) was significantly linked to an SST or SST12 impairment, justifying the no need to a psychophysical screening test to take social distancing and barrier measures in case of acute olfactory disorders in COVID-19 pandemic times. Even if SST-12 has been recently evaluated in a single-use “filter paper” manner^(38)^, 7% of hyposmic patients could be missed with such a test. This is especially true since such a screening test poses a contamination risk to the examiner. However, at a distance from acute infection, SST-12 could be helpful to screen post COVID-19 olfactory disorders. In case of a complaining person, as our results suggest, a complete psychophysical olfactory test might be directly performed as an olfactory complain is highly correlated with an impaired SST and SST-12. But in case of a non-complaining, or olfactory impairment unaware, post-COVID-19 patient, SST-12 could avoid negative consequences of unknown olfactory disorders, especially quality of life^(5,39–41)^ and metabolic impairments. Indeed, 45,5 and 20,5% (n=44) of post COVID-19 patients increased respectively their daily diet salt and sugar intake. As previously published, salt and sugar intake increase concerned near 30% of COVID-19 patients^(15)^, especially young women. Our results suggest that theses bad diet habits could concerned in fact olfactory impaired post-COVID-19 patients, specifically anosmics ones (SST-12≤6) being deprived of their original food tastes and trying to enhance it whatever the way. Interestingly, there is no significant relation, otherwise only with the SST-12 score, between the risk of bad diet habits and subjective olfactory complain, underlining the benefits of using SST-12. The sugar intake is also concerned as COVID-19 could basically raise blood glucose and HbA1c levels^(42)^ which has to be monitored after hospital discharge. It’s a major public health concern as post-COVID-19 olfaction disorders recovery time is still uncertain and long term salt and sugar intake could increase respectively blood pressure^(43)^ and type 2 diabetes^(44)^ onset and so, cardiovascular risk. In case of SST-12 screened anosmia, a not to change daily use of salt and sugar advice must be added to the patient consultation.

Despite these interesting results, this study suffers from some limitations. The main limitation concerns the small cohort of 54 patients, with no follow up reported, who spontaneously consulted our university hospital, which represents the risk of a recruitment bias. The small sample size may have contributed to a limited strength of correlations (rho(32) MAX = 0,49), and therefore our results cannot be directly generalized to all patients with a post-covid olfactory disorder and could be verified in a larger prospective cohort study.

## CONCLUSION

The SST 12 is an olfactory psychophysical test suitable for screening an olfactory sequelae post COVID-19. It makes it possible to highlight the subjective complaint of patients on the odor’s identification. Its use in the context of screening for a long olfactory covid could be used for the implementation of personalized management of olfactory disorders and the prevention of psychological and metabolic consequences adding to the impairment of quality of life.

## Data Availability

The data that support the findings of this study are available from the corresponding author, [CV], upon reasonable request.

## REFERENCES

1. Gerkin RC, Ohla K, Veldhuizen MG, et al. Recent Smell Loss Is the Best Predictor of COVID-19 Among Individuals With Recent Respiratory Symptoms. Chem Senses. 2021;46(December 2020):1–12.

2. Gane SB, Kelly C, Hopkins C. Isolated Sudden Onset Anosmia in COVID-19 Infection. A Novel Syndrome? Rhinol J. 2020;58(3):299–301.

3. Carrillo-Larco RM, Altez-Fernandez C. Anosmia and dysgeusia in COVID-19: A systematic review. Wellcome Open Res. 2020;5:94.

4. Kanjanaumporn J, Aeumjaturapat S, Snidvongs K, Seresirikachorn K, Chusakul S. Smell and taste dysfunction in patients with SARS-CoV-2 infection: A review of epidemiology, pathogenesis, prognosis, and treatment options. Asian Pacific J Allergy Immunol. 2020;69–77.

5. Vandersteen C, Payne M, Dumas L-E, et al. Persistent olfactory complaints after COVID-19: a new interpretation of the psychophysical olfactory scores. Rhinol Online. 2021;4(14):66–72.

6. Hur K, Choi JS, Zheng M, Shen J, Wrobel B. Association of alterations in smell and taste with depression in older adults. Laryngoscope Investig Otolaryngol. 2018;3(2):94–9.

7. Kohli P, Soler ZM, Nguyen SA, Muus JS, Schlosser RJ. The Association Between Olfaction and Depression: A Systematic Review. Chem Senses. 2016;41(6):479–86.

8. Croy I, Nordin S, Hummel T. Olfactory disorders and quality of life-an updated review. Chem Senses. 2014;39(3):185–94.

9. Nordin S. Sensory perception of food and ageing. In: Food for the Ageing Population. Elsevier; 2009. p. 73–94.

10. Valsamidis K, Printza A, Constantinidis J, Triaridis S. The Impact of Olfactory Dysfunction on the Psychological Status and Quality of Life of Patients with Nasal Obstruction and Septal Deviation. Int Arch Otorhinolaryngol. 2020;24(02):e237–46.

11. Schablitzky S, Pause BM. Sadness might isolate you in a non-smelling world: olfactory perception and depression. Front Psychol. 2014;5(FEB).

12. Nordin S, Brämerson A. Complaints of olfactory disorders: epidemiology, assessment and clinical implications. Curr Opin Allergy Clin Immunol. 2008;8(1):10–5.

13. Ahmedy F, Mazlan M, Danaee M, Abu Bakar MZ. Post-traumatic brain injury olfactory dysfunction: factors influencing quality of life. Eur Arch Oto-Rhino-Laryngology. 2020;277(5):1343–51.

14. Hopkins C, Surda P, Vaira LA, et al. Six month follow-up of self-reported loss of smell during the COVID-19 pandemic. Rhinol J. 2020;(11):0–0.

15. Rolland B, Haesebaert F, Zante E, Benyamina A, Haesebaert J, Franck N. Global Changes and Factors of Increase in Caloric/Salty Food Intake, Screen Use, and Substance Use During the Early COVID-19 Containment Phase in the General Population in France: Survey Study. JMIR Public Heal Surveill. 2020;6(3):e19630.

16. Boscolo P, Francesco R, Jerry G, et al. Self - reported smell and taste recovery in coronavirus disease 2019 patients: a one - year prospective study. Eur Arch Oto- Rhino-Laryngology. 2021;(0123456789).

17. Doty RL. Office Procedures for Quantitative Assessment of Olfactory Function. Am J Rhinol. 2007;21(4):460–73.

18. Su B, Bleier B, Wei Y, Wu D. Clinical Implications of Psychophysical Olfactory Testing: Assessment, Diagnosis, and Treatment Outcome. Front Neurosci. 2021;15(March):1– 12.

19. Hummel T, Whitcroft KL, Andrews P, et al. Position paper on olfactory dysfunction. Rhinol J. 2017;54(26):1–30.

20. Agyeman AA, Chin KL, Landersdorfer CB, Liew D, Ofori-Asenso R. Smell and Taste Dysfunction in Patients With COVID-19: A Systematic Review and Meta-analysis. Mayo Clin Proc. 2020;95(8):1621–31.

21. Oleszkiewicz A, Schriever VA, Croy I, Hähner A, Hummel T. Updated Sniffin’ Sticks normative data based on an extended sample of 9139 subjects. Eur Arch Oto-Rhino- Laryngology. 2019;276(3):719–28.

22. Sorokowska A, Schriever VA, Gudziol V, et al. Changes of olfactory abilities in relation to age: odor identification in more than 1400 people aged 4 to 80 years. Eur Arch Oto- Rhino-Laryngology. 2015;272(8):1937–44.

23. Whitcroft KL, Cuevas M, Haehner A, Hummel T. Patterns of olfactory impairment reflect underlying disease etiology. Laryngoscope. 2017;127(2):291–5.

24. Gudziol V, Lötsch J, Hähner A, Zahnert T, Hummel T. Clinical significance of results from olfactory testing. Laryngoscope. 2006;116(10):1858–63.

25. Steinbach S, Hummel T, Böhner C, et al. Qualitative and Quantitative Assessment of Taste and Smell Changes in Patients Undergoing Chemotherapy for Breast Cancer or Gynecologic Malignancies. J Clin Oncol. 2009;27(11):1899–905.

26. Hummel T, Sekinger B, Wolf SR, Pauli E, Kobal G. ‘Sniffin’ Sticks’: Olfactory Performance Assessed by the Combined Testing of Odour Identification, Odor Discrimination and Olfactory Threshold. Chem Senses. 1997;22(1):39–52.

27. Doty RL, Marcus A, William Lee W. Development of the 12-Item Cross-Cultural Smell Identification Test(CC-SIT). Laryngoscope. 1996;106(3):353–6.

28. Jackman AH, Doty RL. Utility of a Three-Item Smell Identification Test in Detecting Olfactory Dysfunction. Laryngoscope. 2005;115(12):2209–12.

29. Allis TJ, Leopold DA. Smell and Taste Disorders. Facial Plast Surg Clin North Am. 2012;20(1):93–111.

30. Rumeau C, Nguyen DT, Jankowski R. Comment tester l’olfaction avec le Sniffin’ Sticks test®. Ann françaises d’Oto-rhino-laryngologie Pathol Cervico-faciale. 2016;133(3):183–6.

31. Hummel T, Rosenheim K, Konnerth C-G, Kobal G. Screening of Olfactory Function with a Four-Minute Odor Identification Test: Reliability, Normative Data, and Investigations in Patients with Olfactory Loss. Ann Otol Rhinol Laryngol. 2001;110(10):976–81.

32. Lechien JR, Chiesa-Estomba CM, Place S, et al. Clinical and epidemiological characteristics of 1420 European patients with mild-to-moderate coronavirus disease 2019. J Intern Med. 2020;288(3):335–44.

33. Lechien JR, Cabaraux P, Chiesa-Estomba CM, et al. Objective olfactory evaluation of self-reported loss of smell in a case series of 86 COVID-19 patients. Head Neck. 2020;42(7):1583–90.

34. Lechien JR, Hopkins C, Saussez S. Sniffing out the evidence; It’s now time for public health bodies recognize the link between COVID-19 and smell and taste disturbance. Rhinol J. 2020;10(7):0–0.

35. Hummel T, Pfetzing U, Lötsch J. A short olfactory test based on the identification of three odors. J Neurol. 2010;257(8):1316–21.

36. Sorokowska A, Oleszkiewicz A, Minovi A, Konnerth CG, Hummel T. Fast Screening of Olfactory Function Using the Q-Sticks Test. ORL. 2019;81(5–6):245–51.

37. Lechien JR, Cabaraux P, Chiesa-Estomba CM, et al. Psychophysical Olfactory Tests and Detection of COVID-19 in Patients With Sudden Onset Olfactory Dysfunction: A Prospective Study. Ear, Nose Throat J. 2020;99(9):579–83.

38. Wirkner K, Hinz A, Loeffler M, Engel C. Sniffin’ Sticks Screening 12 test: Presentation of odours on filter paper improves the recognition rate. Rhinol J. 2021;0–0.

39. Liu DT, Besser G, Prem B, et al. Self-perceived Taste and Flavor Perception: Associations With Quality of Life in Patients With Olfactory Loss. Otolaryngol Neck Surg. 2020;019459982096524.

40. Elkholi SMA, Abdelwahab MK, Abdelhafeez M. Impact of the smell loss on the quality of life and adopted coping strategies in COVID-19 patients. Eur Arch Oto-Rhino- Laryngology. 2021;(0123456789).

41. Coelho DH, Reiter ER, Budd SG, Shin Y, Kons ZA, Costanzo RM. Quality of life and safety impact of COVID-19 associated smell and taste disturbances. Am J Otolaryngol. 2021;42(4):103001.

42. Chen J, Wu C, Wang X, Yu J, Sun Z. The Impact of COVID-19 on Blood Glucose: A Systematic Review and Meta-Analysis. Front Endocrinol (Lausanne). 2020;11(October):1–8.

43. Graudal NA, Hubeck-Graudal T, Jurgens G. Effects of low sodium diet versus high sodium diet on blood pressure, renin, aldosterone, catecholamines, cholesterol, and triglyceride. Cochrane Database Syst Rev. 2017;2017(4).

44. Lean MEJ, Te Morenga L. Sugar and type 2 diabetes. Br Med Bull. 2016;120(1):43–53.

